# Piloting the Clinical Value of Wearable Cardiorespiratory Monitoring for People with Cystic Fibrosis

**DOI:** 10.1101/2020.07.27.20151589

**Authors:** Benjamin Vandendriessche, Bertold Van den Bergh, Valerie Storms, James F. Chmiel, Erica A. Roesch

## Abstract

**Introduction:** Cystic Fibrosis (CF) is a disease without a primary cure that requires lifelong care and is characterized by pulmonary exacerbations (PEx). Wearable devices could provide a way for long-term monitoring of disease progression and early signs of PEx to intervene as early as possible, thereby improving long-term outcomes.

**Methods:** In-hospital feasibility study (n = 26) to 1) assess the ability of Byteflies Sensor Dot to collect relevant cardiorespiratory data in people with CF and its compatibility with clinical workflows, 2) identify candidate digital biomarkers, and 3) collect user feedback from patients and healthcare providers.

**Results:** Collected sample-level biopotential, bioimpedance and actigraphy data were of high quality. Sensor Dot heart rate (HR) correlated with hospital HR, whereas respiratory rate (RR) did not. HR and RR were associated with CF severity, and HR and coughing with PEx. Willingness to use the device was very high with CF patients and study coordinators considered the device easy-to-use.

**Conclusion:** Determining if a wearable is fit-for-purpose is a long and multidisciplinary process that requires involvement from all stakeholders as early as possible in the development process. Our pilot identified interesting correlations between cardiorespiratory parameters as measured by the wearable, and CF severity and PEx. Together with the usability data, this will inform the next steps in the clinical development process.

## Introduction

Cystic Fibrosis (CF) is an autosomal recessive disorder caused by mutations in the CFTR gene that affects about 70,000 to 100,000 people worldwide [1]. No primary cure is available. The associated symptoms mainly affect the respiratory and digestive systems, and are complex, progressive, and require lifelong multifactorial care [2]. Significant improvements in early detection via newborn screening tests and targeted modulator therapies have had a positive impact on life expectancy, but frequent, personalized, and aggressive interventions are still needed [2,3].

Currently, the Cystic Fibrosis Foundation recommends that people ages 6 years and older visit their care center at least 4 times and perform at least 2 pulmonary function tests (PFTs) per year [4]. Between clinic visits, little to no objective monitoring is available. Clinicians are often reliant on an individual’s recognition and self-reporting of symptoms. However, patient reported outcomes (PROs) specific for pulmonary exacerbations (PEx) in CF are known to be inaccurate and unreliable [5]. User-friendly and clinically validated wearable devices could fill this void by continuously monitoring affected organ systems to generate a more holistic view into the patient’s health status. This includes assessing the influence of behavioral patterns and lifestyle changes, treatments, and PROs. Because early detection and treatment of PEx in CF is so important to reduce cumulative damage to lung tissue and subsequent morbidity and mortality [6], cardiorespiratory monitoring with a wearable is an interesting and unexplored strategy to continue to improve the standard of care for people with CF.

Qualification of a digital biomarker for PEx in CF would require appropriate validation studies [7], but before going down that route, it is important to understand the disease course and care management regimens, the potential context(s) of use, and the willingness of a person with CF to use the tool. Medical wearables require rigorous verification, analytical, and clinical validation (V3), which ideally incorporates feedback from all stakeholders continuously and as early as possible in the process [8]. Therefore, we conducted a pilot study with 3 main objectives: 1) test Byteflies Sensor Dot, a prototype wearable that continuously collects cardiorespiratory data to evaluate how it could be incorporated into clinical workflows for the management of people with CF, while the device is still in a modifiable state to become fit-for-purpose, 2) evaluate the collected data for clinically useful trends and other patterns that could become candidate digital biomarkers, and 3) collect user feedback from healthcare professionals and patients. This pilot study was run in the relatively controlled environment of the CF clinic and hospital ward in anticipation of longer-term home monitoring.

## Materials and Methods

### Study Design

Outpatients and inpatients with Cystic Fibrosis (CF) at the Rainbow Babies and Children’s Hospital of University Hospitals Cleveland Medical Center (UHCMC) were recruited to wear a Sensor Dot, a wearable that records cardiorespiratory and activity parameters for the full or partial duration of their hospital stay. Inclusion criteria were: ≥ 18 years old and confirmed CF diagnosis as determined by a sweat chloride ≥ 60 mmol/L or the presence of two known disease-causing mutations. Exclusion criteria were: 1) inability to provide written informed consent, 2) a known allergy to any of the used medical adhesives, and 3) presence of any type of electronic implanted medical device. Patients were enrolled between December 2018 and June 2019. All experiments were approved by the Institutional Review Board of UHCMC and written informed consent was obtained for all study participants. Selected study data were collected and managed using REDCap electronic data capture tools, hosted at UHCMC [9].

### Data Collection

Physiologic data were collected with a prototype version of Sensor Dot, a small wearable that can record biopotential (ExG) and bioimpedance (bioZ) signals, and actigraphy via a tri-axial accelerometer (ACC). After explaining the study purpose, the Sensor Dot and electrodes were applied to the subject in an approximate ECG Lead 2 configuration (Fig. 1), after which the subject continued with the visit protocol or hospitalization as indicated. The Sensor Dots were configured to collect 1 channel of electrocardiography (ECG) at 125 Hz, 1 channel of respiration (RES) at 125 Hz, and ACC at 50 Hz per channel. Authorized UHCMC and Byteflies staff could access the data over a secure web interface. Study participants were asked to fill out a user experience survey in REDCap at the conclusion of their participation. Relevant demographic (age, sex), PFT, and vital sign (temperature, heart rate (HR), respiratory rate (RR), blood pressure (BP), and oxygen saturation (SpO_2_)) data were extracted from the electronic health record and added to the participant’s REDCap record. Sensor Dots were cleaned before using them on another study participant.

**Fig. 1.**
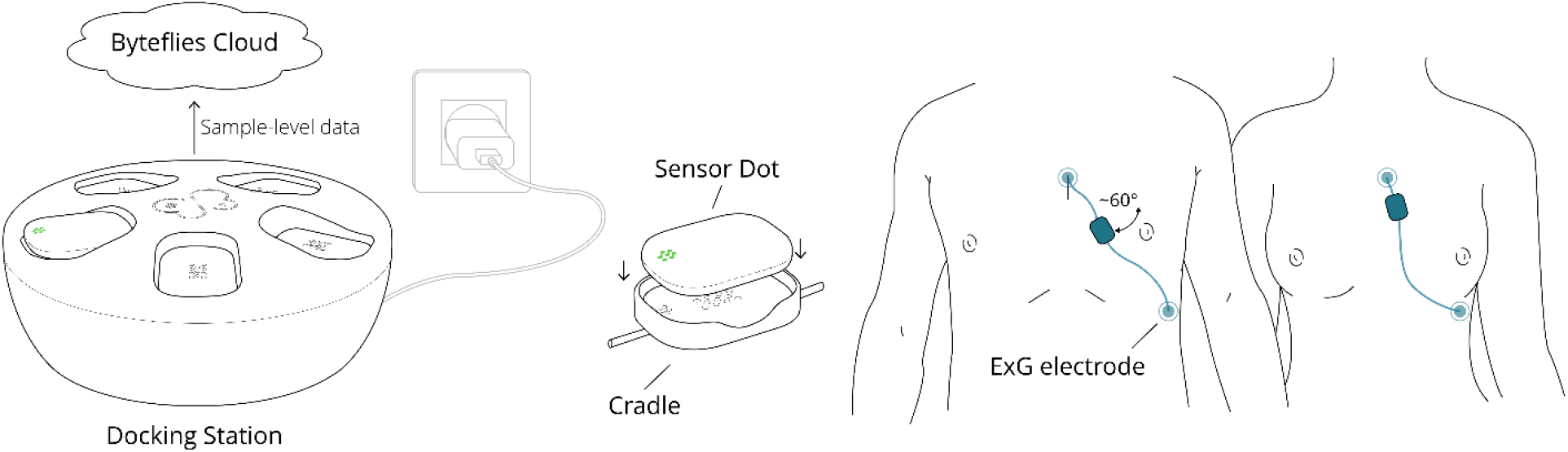
The Byteflies Sensor Dot System. (from left-to-right) Sensor Dots (34×25×8 mm, 6 g) are idle in the Docking Station. To start a recording, a Sensor Dot is placed in a cradle which provides an interface to commercially available ExG electrodes for measuring ECG and RES. The cradle is attached to the skin using a double-sided medical adhesive. To record cardiorespiratory parameters, the negative lead is placed on the sternum, approximately 4 cm below the clavicle, and the positive lead is placed near intercostal space 6-7 on the left side the thorax and distal to the heart’s apex, at an angle of approximately 60 degrees relative to the shoulders. As long as the wearable device was on the patient, sample-level (raw) data were logged on the onboard memory. To complete a measurement, the study coordinator removed the Sensor Dot and electrodes from the patient and the Sensor Dot was placed in the Docking Station to charge its battery and upload the recorded physiologic data with a patient identifier to the HIPAA-compliant Byteflies Cloud.

### Data Processing

All sample-level and processed data described below were visually examined to evaluate the output of the algorithms used for data exploration.

#### 1. Electrocardiogram (ECG)

ECG-parameters were calculated via a proprietary Byteflies algorithm. Briefly, 1) ECG waveforms were preprocessed to remove baseline noise and exclude areas of low quality data, 2) a convolutional neural network (CNN) trained on approximately 36,000 h of ECG data then identified the ECG R-peaks, and 3) post-processing steps examined the interbeat-intervals (IBI) for artifacts (technical outliers) and ectopic beats (physiologic outliers), and corrected them via linear-spline interpolation if appropriate. The resulting IBI time series was used to calculate HR by averaging over 10 sec windows. The quality of R-peak detection was visually evaluated for each recording based on their IBI Poincaré plot [10].

#### 2. Respiration (RES)

The bioZ signal modulates with the respiratory volume and chest movement, and is therefore a promising signal for long-term, passive, and unobtrusive respiratory monitoring [11]. Respiratory parameters were calculated using the *scipy* Python library. Briefly, 1) the RES signal was bandpass filtered between 0.1 and 1 Hz, and 2) respiratory cycles were detected using peak detection set to identify cycles that are spaced apart at least 1.5 sec, and with a peak width at 80% of the peak height of at least 1 sec. The resulting respiratory cycles were used to calculate RR and the amplitude of respiratory cycles (RES_AMP_) by taking the median for each 10 sec window. The quality of respiratory cycle detection was evaluated for each recording by comparing RR_RES_ to RR_ACC_ (see Actigraphy).

#### 3. Actigraphy (ACC)

Three parameters were derived from the ACC signal: 1) an activity index (AI), as outlined in [12], which provides a summarized representation of the sample-level ACC data, 2) RR, and 3) a rudimentary measure for coughs. The AI with a correction for systematic noise was calculated for each 1 sec window [12], followed by taking the median AI for each 10 sec window. Respiratory parameters were calculated for the ACC signal using the standard *scipy* Python library: 1) the z-axis ACC, which is positioned perpendicular to the chest and thus the main direction of respiration-related chest wall motion, was low-pass filtered at 0.04 Hz, and 2) respiratory cycles and RR were calculated as described before. If RR_RES_ diverged from RR_ACC_ by more than 15%, the signals were visually evaluated. Finally, events that are likely coughs were detected by: 1) high-pass filtering the z-axis of the ACC signal at 0.4 Hz (ACC-Z_HP_), 2) taking the power of the high-passed signal (ACC-Z_HP_^2^), 3) slicing ACC-Z_HP_^2^ based on the respiratory cycles derived from the RES signal since a cough will lead to an RES inflection, and 4) taking the area-under-the-curve (AUC) of ACC-Z_HP_^2^ for each slice. For this proof-of-concept implementation, a threshold was set for each study participant based on visual inspection of the data. Specifically, the RES signal was examined together with the AUC results and the threshold was set to include events that resembled a cough on the RES signal (corroborated by the ACC-Z_HP_^2^ signal derivation).

#### 4. Comparison to Hospital Vital Signs

As the length of Sensor Dot data for study participants varied widely (1-23 h), it would be difficult to make direct comparisons to other discrete measurements. Therefore, REST and ACTIVE epochs were defined for each recording: 1) REST was defined as the average of 3 epochs of 5 min each during restful wake where AI < 1, and 2) ACTIVE was defined as the average of 2 epochs of 5 min each during activity where AI was highest for that subject. δHR is defined as HR_ACTIVE_ -HR_REST_, and similar for other vital signs. No sleep phases were used as they were only available for some of the inpatients. Since participants were not asked to complete specific physical tasks, the ACTIVE and REST groups exhibited large heterogeneity which could be somewhat reflective of real-world conditions, although confounded by the hospital environment and variability in recording length.

HR, BP, SpO_2_, and body temperature were measured using a Welch Allyn Connex Vital Signs Monitor or the GE Carescape V100 Vital Signs Monitor. RR was manually counted. Hospital-measured HR and RR were compared to the same parameters under REST conditions for Sensor Dot data.

#### 5. Comparison to PFT

Calibration, quality control of equipment, and spirometry were performed according to ATS guidelines [13]. Percent predicted values were calculated using the Global Lung Function Initiative (GLI) values [14]. Participants were divided into 3 severity categories based on baseline PFT results: mild (FEV_1_≥ 70%), moderate (69% > FEV_1_ ≥ 40%), and severe (FEV_1_ < 40%). HR, RR, and RES_AMP_ derived from Sensor Dot data for the REST and ACTIVE epochs, and the total length of each recording, were examined as a function of severity.

#### 6. User Survey

User surveys probed for participant’s experience with the Sensor Dot prototype and its perceived value for managing the individual’s condition. Categorized results were aggregated. Study participants were also asked to estimate the number of times they coughed while wearing the device. Coughs counted by Sensor Dot were compared to self-reported cough counts. Study coordinators were interviewed about their experience with the device.

### Statistics

All analysis of waveform data was done in Python. All comparative analyses of Sensor Dot versus hospital-measured parameters were done with GraphPad Prism 8.02. Linear regression was performed for the vital sign comparisons and R^2^ values are reported. One-way ANOVA (95% confidence interval (CI)) was used for comparison of age, PFT, and hospital vital signs to PFT severity categories with Tukey’s correction for multiple comparisons. Two-way ANOVA (95% CI) was used for all other comparisons to PFT severity categories and patient types with Tukey’s correction for multiple comparisons. An unpaired t-test (95% CI) was used to compare RES amplitude results between patient types. Reported values are mean ± SD unless otherwise indicated.

## Results

### Study Cohort and Clinical Workflow

A total of 26 subjects participated in the study (46% inpatients). The mean cohort age was 31.9 ± 12.3 years (65.4% male). Subjects were divided into mild, moderate, and severe categories (Table 1). A trend for increased age and decreased SpO_2_ was observed in the higher severity categories. There were no differences in HR, RR, BP, or body temperature between severity categories.

**Table 1.** Demographics, PFT results, and basic vital signs collected as part of routine care procedures. Participants were divided into 3 severity categories based on baseline PFT results: mild (FEV_1_≥ 70%), moderate (69% > FEV_1_ ≥ 40%), and severe (FEV_1_ < 40%). FEV: forced expiratory volume, FVC: forced vital capacity, FEF: forced expiratory flow, pp: percent predicted, BPM: beats-per-minute, BrPM: breaths-per-minute, SYS: systole, DIAS: diastole. *, p < 0.05; **, p < 0.01; ***, p < 0.001; ****, p < 0.0001; ns: not-significant; p-values are indicated for borderline ns results.

|  | Mild (A) | Moderate (B) | Severe (C) | Statistics |
| --- | --- | --- | --- | --- |
| **Subjects** (n (%)) | 10 (38%) | 9 (35%) | 7 (27%) | NA |
| **Inpatients** (n (%)) | 4 (44%) | 3 (33%) | 5 (71%) | NA |
| **Age** (mean ± SD) | 25.4 ± 6.5 | 35.7 ± 11.3 | 38.0 ± 16.0 | A-B: ns; B-C: ns; A-C: 0.08 |
| **Sex** (M / F) | 70% / 30% | 67% / 34% | 43% / 57% | NA |
| **FEV<sub>1</sub>** (L (pp)) | 3.18 ± 0.62<br>(83.3 ± 9.2) | 2.12 ± 0.43<br>(56.9 ± 9.9) | 1.25 ± 0.23<br>(34.0 ± 3.3) | A-B: ****; B-C: ****; A-C: **** |
| **FVC** (L (pp)) | 4.30 ± 1.00<br>(95.0 ± 12.0) | 3.58 ± 0.92<br>(77.9 ± 14.1) | 2.32 ± 0.60<br>(51.6 ± 8.3) | A-B: *; B-C: ***; A-C: **** |
| **FEF 25-75** (L (pp)) | 2.43 ± 0.60<br>(58.8 ± 16.4) | 1.03 ± 0.37<br>(26.9 ± 9.7) | 0.50 ± 0.11<br>(13.6 ± 2.9) | A-B: ****; B-C: ns; A-C: **** |
| **FEV<sub>1</sub> / FVC** | 74.6 ± 5.9 | 61.0 ± 10.2 | 54.6 ± 5.3 | A-B: ** B-C: ns; A-C: **** |
| **HR** (BPM) | 87.1 ± 16.8 | 83.4 ± 10.8 | 87.3 ± 9.3 | A-B: ns; B-C: ns; A-C: ns |
| **RR** (BrPM) | 19.0 ± 3.0 | 18.3 ± 1.4 | 19.6 ± 1.7 | A-B: ns; B-C: ns; A-C: ns |
| **SpO<sub>2</sub>** (%) | 97.0 ± 1.6 | 96.6 ± 1.0 | 95.4 ± 1.7 | A-B: ns; B-C: ns; A-C: 0.09 |
| **Temperature** (°C) | 36.4 ± 0.3 | 36.7 ± 0.2 | 36.7 ± 0.1 | A-B: *; B-C: ns; A-C: * |
| **BP** (SYS (DIAS)) | 119.3 ± 6.8<br>(70.9 ± 18.3) | 119.7 ± 10.4<br>(73.0 ± 9.5) | 118.4 ± 15.2<br>(73.9 ± 4.5) | A-B: ns; B-C: ns; A-C: ns |

### Data Quality

Waveform data and the output generated by data exploration algorithms were visually and algorithmically evaluated. Approximately 135 h of data were recorded, with individual recordings ranging from 1-23 h. For 3 recordings (11.5%) the bioZ signal was not recovered due to a firmware bug. For those recordings, only RR_ACC_ was used. For another 4 subjects (15.3%), only partial recordings contained usable data due to bad ECG electrode contact (6.9% (557 min) data loss). After removing this low-quality data, HR was of satisfactory quality for all remaining data (7543 min). Physical motion that involves the thorax can potentially distort the RR_RES_, RES_AMP_, and RR_ACC_ parameters but the RES and ACC signals can also corroborate each other [11]. When examining summarized RR_RES_ and RR_ACC_, they track each other closely, although RR_ACC_ has a higher SD compared to RR_RES_ (Fig. 2, middle panel). The associated averaged RR_RES_ and RR_ACC_for both REST epochs as well as the total recording length are moderately correlated (Fig. 3C, R^2^ = 0.44 and 0.55, respectively). The AI derived from ACC was of satisfactory quality for all recorded data as it does not rely on electrodes.

**Fig. 2.**
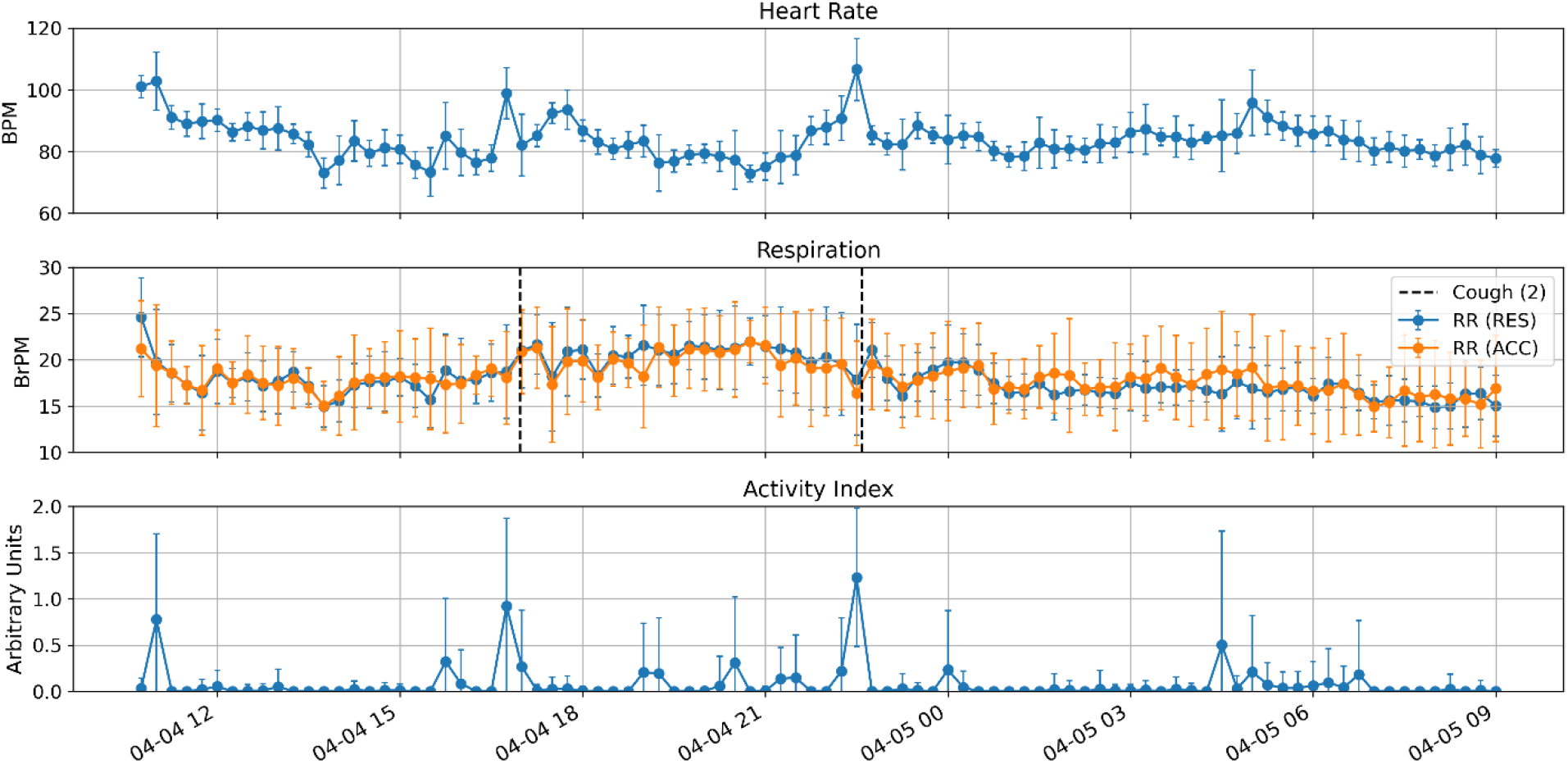
Representative longitudinal (22 h) physiological and activity profile for one study subject. (Top) Heart rate (HR). (Middle) Respiration Rate (RR) derived from the RES (blue) and ACC (orange) signal, as well as cough events (dashed vertical marker). (Bottom) Activity Index (AI). All data are binned per 15 min and means ± SD.

**Fig. 3.**
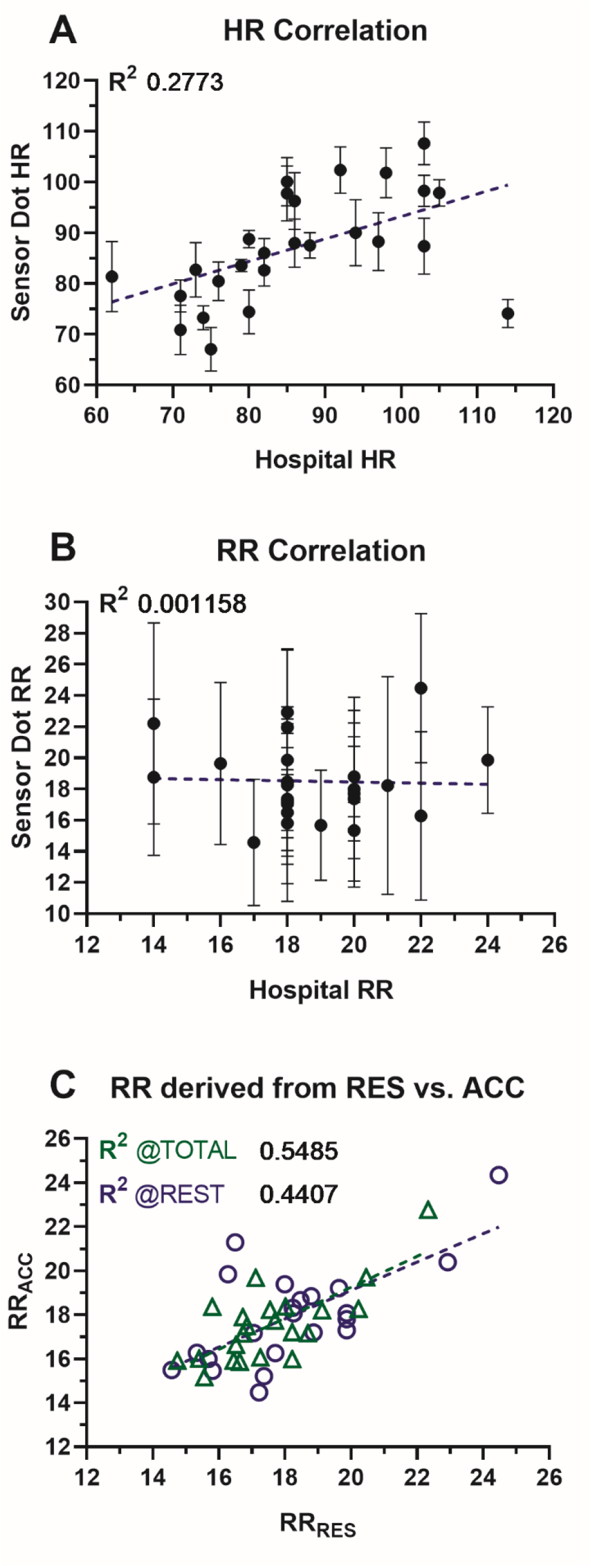
Correlation of hospital to Sensor Dot vital signs. (A) Linear regression (blue, R^2^: 0.28) of HR as derived from the ECG signal and hospital equipment. Error bars are SD. (B) Linear regression (blue, R^2^: 0.001) of RR as derived from the RES signal and manually counted in the hospital. Error bars are SD. Linear regression of RR_RES_ and RR_ACC_ under REST (blue R^2^: 0.44) conditions, and for all (TOTAL) recorded data (green, R^2^: 0.55).

### Clinical Insights

For each study subject, the HR, RR_RES_, RR_ACC_, cough events, and AI were calculated (Fig. 2). The HR and RR spot checks performed in the clinic or hospital were compared to the average Sensor Dot HR and RR during the REST epochs. HR was weakly correlated (Fig. 3A, R^2^ = 0.28), which is not surprising considering a brief hospital measurement is compared to one based on three 5 min epochs, arbitrarily distributed over the recording. Nevertheless, it indicates some of the resting HR patterns are preserved, at least within a single day. Although the prototype version of Sensor Dot at the time of this study did not yet undergo full analytical validation against a predicate device, we know the HR results are correct because: 1) the device underwent verification testing according to international ECG standards, and 2) the sample-level ECG data and R-peak annotations are available and were visually inspected. The RR hospital results (based on manual counting) were not correlated with Sensor Dot RR (Fig. 3B, R^2^ = 0.001). Like the ECG case, we can visually inspect the sample-level RES data and respiratory cycle annotations. However, as a respiratory signal is typically less regular than an ECG signal, it is not possible to make a similar claim with as much certainty as for the ECG case without comparison to a well-validated reference device.

HR and RR for REST, ACTIVE, and total recording length were examined as a function of the CF severity (mild, moderate, severe). Compared to the mild group, HR was significantly increased for the moderate and severe groups under ACTIVE conditions, and there was a similar trend under REST conditions (Fig. 4A1). δHR was significantly increased in the inpatient versus outpatient groups (Fig. 4A2). RR was significantly increased in the severe compared to the mild and moderate groups under REST conditions, and a similar trend was observed for the ACTIVE and total recording length epochs (Fig. 4B1). No meaningful patterns were identified for δRR or RES_AMP_ for the severity categories. When comparing RES_AMP_ across inpatients and outpatients, a non-significant increasing trend was observed (Fig. 4B2). Finally, the number of self-reported coughs were compared to Sensor Dot coughs. No significant differences were found between severity categories (Fig. 4C1), but a higher number of coughs was identified for inpatients, both self-reported and on Sensor Dot (Fig. 4C2). The averaged self-reported and Sensor Dot-derived number of coughs were similar (p > 0.99).

**Fig. 4.**
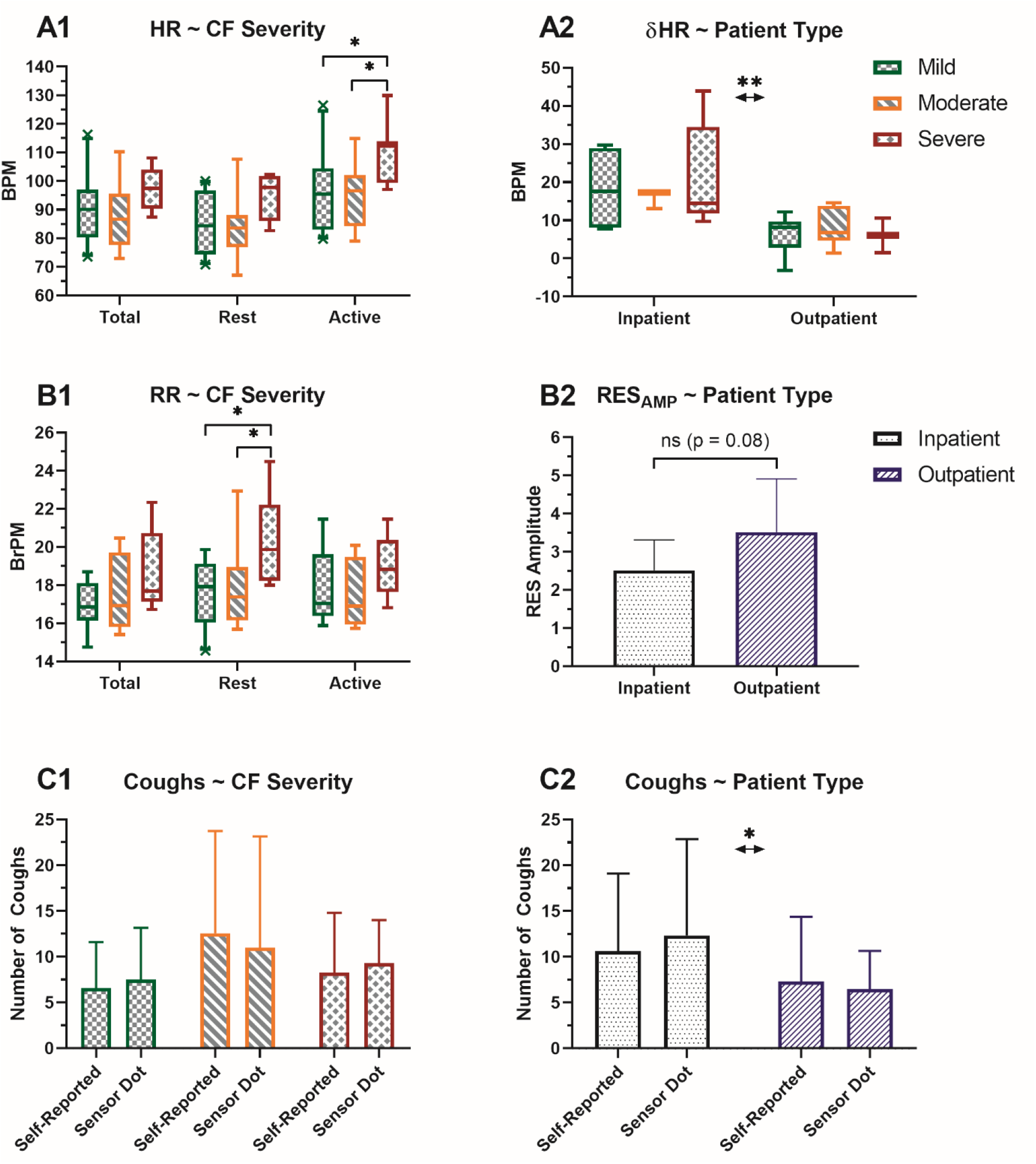
Comparison of Sensor Dot vital signs to the CF severity classification. (A1) HR comparison: Total summarizes the results for all recorded Sensor Dot data; REST and ACTIVE are as defined in Materials & Methods. 10-90 percentile are plotted with means indicated. The “mild vs. severe” and “moderate vs. severe” comparisons in the ACTIVE group are p = 0.0260 and p = 0.0177, respectively. (A2) δHR (HR_ACTIVE_ -HR_REST_) compared across severity categories and patient type. The patient type effect (inpatient vs. outpatient) is p = 0.0046 (**). (B1) RR comparison identical to panel A1. The “mild vs. severe” and “moderate vs. severe” comparisons in the REST group are p = 0.0124 and p = 0.0268, respectively. (B2) Mean RES_AMP_ for inpatients and outpatients. A trend (p = 0.0822) for decreased RES_AMP_ in the inpatient group was observed. (C1) Mean number of recorded cough events on Sensor Dot vs. self-reported. Coughs were self-reported in brackets: < 5, 5-10, 11-20, 21-30, > 30. (C2) Mean number of recorded cough events for inpatients vs. outpatients. The patient type effect is p = 0.0429 (*). *p < 0.05; **p < 0.01, error bars are SD.

### User Feedback

Each study participant was asked to complete a survey after wearing Sensor Dot to assess comfort, willingness to wear the device in a clinical setting and at home, acceptable wear time, and user experience. Results were overall positive. All participants who completed the survey scored the device as comfortable (Fig. 5A). Most study participants indicated that that they would absolutely or probably (81%) wear the device if asked to do so by their physician, with 12% indicating they would maybe wear it (Fig. 5B). Forty-two percent of participants indicated they would be willing to wear the device a couple of days per week, while 8% were willing to wear it during the day only, and another 8% stated they would wear it less than once a week (Fig. 5C). Averaged results from questions assessing the user experience showed that 88% of participants considered their experience very positive to positive and 5% as neutral to negative. Study coordinators found the device easy to set up and attach to the patient. Once instructed, the process took approximately 5 minutes. Because the devices were pre-configured for the study, all other actions were automatic and did not interfere with clinical workflows.

**Fig. 5.**
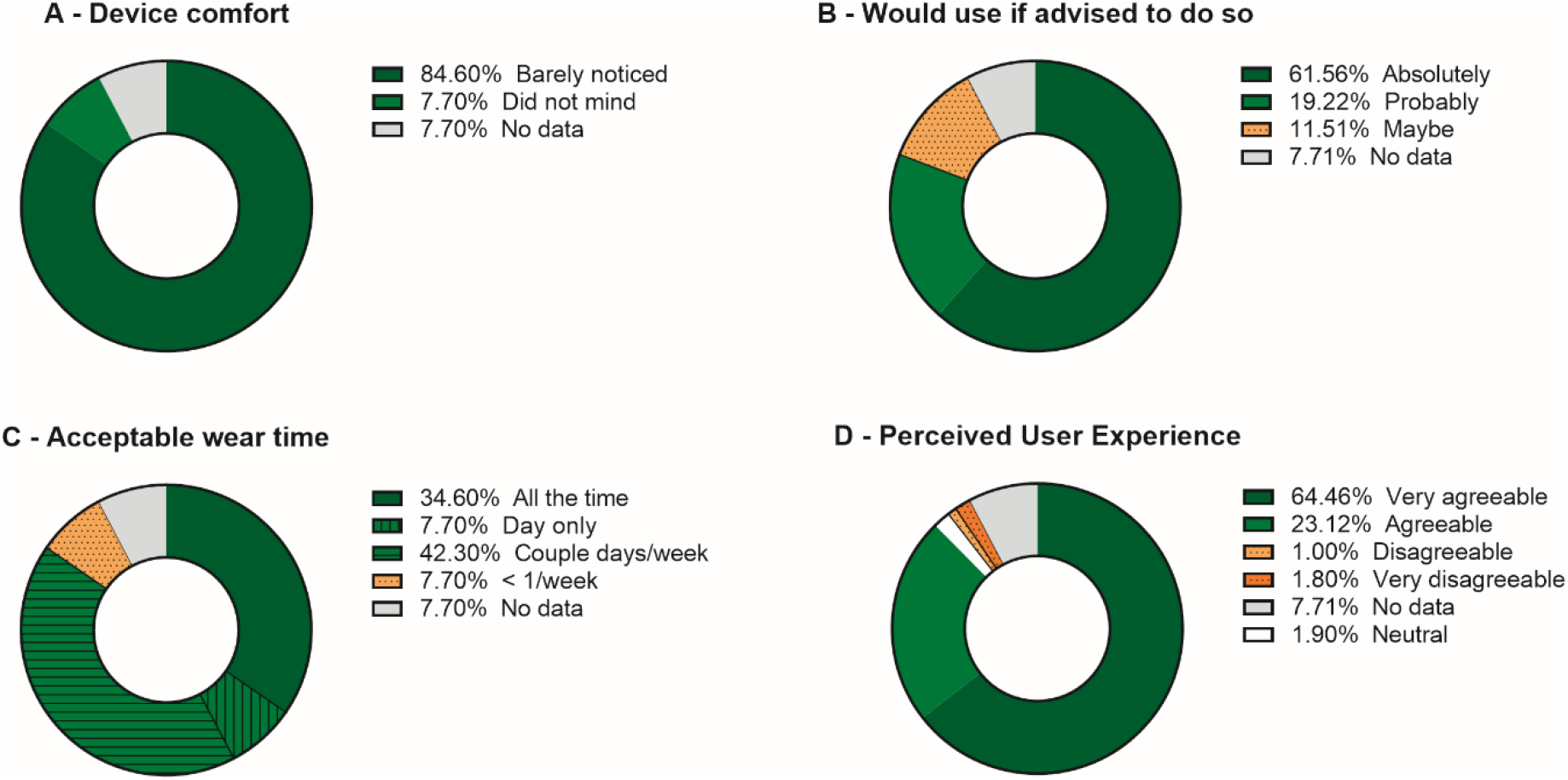
User survey results. 2/26 participants did not complete the survey due to technical difficulties (gray category) (A) “Do you think the device was comfortable to wear?”, options were: “Yes, barely noticed it was there”; “Yes, did not mind it”; “Okay, it was somewhat distracting/annoying”; “No, I consistently noticed it was there”; “No, I would never wear it again”. (B) “Would you be willing to wear a device like this at home if your doctor recommended it?”, options were: “Absolutely”; “Probably”; “Maybe”; “Never”. (C) “How long would you be willing to wear a device like this?”, options were: “All the time”; “Only during the day”; “Only during the night”; A couple of days per week”; “Less than once weekly”. (D) Perceived user experience was aggregated from 5 questions that were graded on a 5-point scale: “I found this device unnecessarily complex”; “I think I would need the support of a technical person to be able to use this device”; “I would imagine that most people would learn to use this device very quickly”; “I found this device very awkward to use”.

## Discussion

We performed a feasibility study to evaluate the use of a cardiorespiratory monitoring wearable, Byteflies Sensor Dot, in the care management of people with CF, in the inpatient and outpatient settings. The wearable has a modular design ensuring that specific components can be iteratively optimized. This is important to ensure it can become fit-for-purpose, while still adhering to verification, analytical and clinical validation (V3) requirements as a clinical care and drug development tool [8]. The collected data were examined *a posteriori* for patterns that might be interesting digital biomarker candidates. Finally, patient and healthcare professional feedback was collected, which served as input information to continue refining Sensor Dot for people with PEx, characteristic for CF and other chronic respiratory conditions. Due to its pilot nature, the study does not provide definitive proof for either of the discussed topics, but rather is a documented step in the development roadmap from general purpose physiologic and behavioral monitoring device towards one with specific clinical indications in CF.

### Study Takeaways

Evaluation of data quality revealed a 7% loss of sample-level data due to bad ECG electrode contact. Combined with the need for people with CF to use Sensor Dot outside the hospital without assistance from a trained person, the use of off the shelf electrodes has since been replaced with an adhesive patch that can be self-administered at home. A larger home study will evaluate the performance of this component.

The primary value of wearables comes from their ability to establish an individual’s physiologic and behavioral baseline, and track deviations from this baseline over various time scales. Individual recordings in this study were at most 23 h long. The goal is to extend this to much longer time frames to create longitudinal profiles that can be augmented with data from other sources, such as home spirometers and PRO tools. The current pilot study is valuable as it guides determination of the eventual context(s) of use early in the development process and takes frequent stakeholder input into account. For instance, will Sensor Dot generate patient value by collecting additional data in between clinic visits, to be presented to their physician during an outpatient visit or admission? Will it evolve into a full telemonitoring application with built-in decision support to warn about impending PEx and allow early intervention? Or, will it become a clinically validated biometric monitoring technology (BioMeT) with a qualified digital biomarker for clinical and research use [7]? The user feedback seems to support all these scenarios, especially if the device can be used frequently but not continuously (see Fig. 5). In all cases, integration with routine clinical workflows will be required to ensure buy-in from healthcare professionals, so starting in that environment is valuable.

Direct comparison to appropriate reference devices is an essential component of any V3 roadmap. In our study, we did not perform a formal analytical validation, but rather integrated Sensor Dot into routine clinical care pathways. We compared the vital signs derived from the wearable to those captured in the hospital. For HR, similar trends were observed but for RR, which is measured via manual counting in the hospital, no correlation with the wearable RR was found. As the results of the RR algorithm were verified visually on the sample-level data, this is likely due to inaccuracies in the manual counting, possibly combined with a change in breathing pattern due to counting awareness. Importantly, because Sensor Dot can sample certain physiologic phenomena with more than one sensor type simultaneously, it is possible to corroborate calculated digital measures using internal reference signals. For instance, bioZ provides a proxy for respiration based on physical chest wall expansion and air volume in the lungs, while ACC provides a proxy based on chest wall motion. This does not negate external analytical validation, but it provides an algorithmic strategy for robust vital sign data collection under real-world conditions. Similarly, a proof-of-concept algorithm for cough detection, based on ACC and bioZ features, seems to correlate well with self-reported coughing, and warrants further dedicated validation work.

Although our study is relatively small in scope, we calculated several digital measures and evaluated their association with CF severity, and whether the patients were inpatient or outpatient. Because inpatients were admitted for a PEx, this group may show differences in certain cardiorespiratory measures that could prove useful to detect PEx early to corroborate unreliable PROs [5]. HR was increased in severe CF, especially during periods of increased physical activity. Interestingly, δHR was significantly increased in the inpatient group compared to outpatients, indicating that the cardiorespiratory effects of a PEx may be captured in the HR response to physical activity. Resting RR was also increased in severe CF. The amplitude of the bioZ signal is not calibrated for lung volume changes but it may nevertheless modulate with changes in respiratory dynamics. A trend for a decreased respiratory amplitude was identified in the inpatients but this was a weak effect that requires further investigation. Similarly, the number of coughs was higher in the inpatient group. As cardiorespiratory fitness is known to correlate with long-term outcomes in CF [15], as does coughing [16], the combination of these factors provide us with a first set of digital measures that may be developed further into one or more qualified digital biomarkers for CF and other chronic respiratory diseases.

## Conclusion

Determining if a BioMeT is fit-for-purpose is a long and multidisciplinary process that requires involvement from all stakeholders as early as possible in the development process. We presented a pilot study that assessed the potential clinical value of a wearable for cardiorespiratory monitoring in CF as an early step in this process. Wearables like Sensor Dot are poised to become part of routine clinical care, and the global COVID-19 pandemic has increased the sense of urgency further, especially for those most vulnerable.

## Data Availability

All data referred to in the manuscript can be requested from the authors.

## Statements

## Acknowledgements

The authors would like to thank the individuals with CF and research coordinators, Brittany Hirth and Cindy Schaefer, who participated in this study.

## Statement of Ethics

All experiments were approved by the Institutional Review Board of UHCMC and performed in accordance with the World Medical Association Declaration of Helsinki. All study participants provided written informed consent.

## Disclosure Statement

BVD and BVB are employees of Byteflies. VS is an employee of Roche. JFC reports grants from the Cystic Fibrosis Foundation (CFF) during the conduct of the study; and grants and personal fees from the CFF, grants from the National Institutes of Health, personal fees from the American Board of Pediatrics, outside the submitted work.

## Funding Sources

Byteflies sponsored the study. Grant support from the US Cystic Fibrosis Foundation for salary support for EAR and JFC and infrastructure support to UHCMC is gratefully acknowledged.

## Author Contributions

BVD, JFC and EAR conceived and designed the study. EAR and BVB coordinated data collection. BVD and EAR analyzed data. BVD, VS, JFC and EAR wrote or critically revised the manuscript.

## Notes

### Clinical Trial

NCT04489186

### Author Declarations

All experiments were approved by the Institutional Review Board of University Hospitals Cleveland Medical Center (UHCMC).

